# Physical health of care-experienced young children in high-income countries: A scoping review

**DOI:** 10.1101/2025.04.15.25325761

**Authors:** Daniel R. R. Bradford, Adam Swift, Mirjam Allik, Alex D. McMahon, Denise Brown

## Abstract

Care-experienced children – also referred to as children in out-of-home care, children in foster care, or looked after children – face additional barriers to good physical health compared to those without care experience. Despite good health in early years being vital to long-term quality of life, there is little research on physical health outcomes in young care-experienced children. This scoping review aimed to collate and review peer-reviewed published literature to identify gaps and inform future research and policy. Standard rigorous scoping review methods were applied. Studies were included if they reported on physical health outcomes affecting children under seven years in high-income countries with care experience. MEDLINE, CINAHL, and Web of Science Core Collection databases were searched. Searches yielded 17,363 results, and 36 articles were included. Studies took place in kinship, foster, residential, and adoptive care settings. Synthesis of results identified poor physical development in terms of height and weight, poor dental health, dermatological conditions, anaemia, and low immunisation rates as substantial health problems among young care-experienced children. However, strong conclusions about the causes and relative prevalence of most conditions could not be drawn. This was often due to a lack of comparator groups, failure to adjust for socioeconomic variables, insufficient reporting about care context, and heterogeneity in study methods. Future work would benefit from relevant comparator groups, clear reporting of participant socioeconomic characteristics and care settings, and limiting focus to specific developmental stages.

## 1 Introduction

Health in the early years has implications for health over the life course [1, 2]. Yet, inequalities in health are apparent from birth and persist across all ages [3–7]. In young children these inequalities are often a result of inadequate provision of the support that they require to thrive. One group of children that are particularly likely to have received inadequate support for their health are care-experienced children – also referred to as children in out-of-home care, children in foster care, or looked after children. These are children who, due to their biological parents being unable to provide adequate care, are moved to alternative caregiving arrangements. Despite the critical importance of achieving good health for young care-experienced children, there is relatively little research about the physical health outcomes which affect these children. The aim of our work was to collate and synthesise the available evidence on this topic, identify gaps in the published literature, enhance understanding of this group’s health needs, and help guide future research and policy directions.

In many countries, legal frameworks exist to safeguard children when parental care is insufficient. Legal guardianship of the child can be transferred partially or completely to other individuals or organisations. This can include kinship care (living with friends or family members other than parents), foster care, residential care, and being adopted. Some countries also have policies that allow children to remain in the parental home while all members of the family receive additional support from social services. Recent estimates of the rate of children currently in care in high-income countries include 12.3 per 1,000 children in Scotland [8], 8.1 in Australia [9], 7.0 in England [10], 5.1 in the USA [11], and 4.0 in Sweden [12]. These values are the rate of children in care on a single day and under-represent the lifetime prevalence of a child having been in care. There is a duty to better understand the health needs of the substantial numbers of young care-experienced children to enable all children to achieve their full health potential.

Care-experienced children have poorer outcomes and experience a higher prevalence for many physical health conditions when compared to children that have never been in care. For example, children in foster care in the USA were more likely to be obese, to have hearing and vision problems, and to be rated as being in poor health in a large nationally-representative survey [13]. Care-experienced individuals are also more likely to die during childhood and live shorter lives [14–16]. Young children in care in Scotland have also been found to be more than twice as likely to need urgent treatment during universal dental health reviews at age five years [17]. This echoes other studies which highlight the issue of poor dental health in care-experienced children [18–20]. Other health conditions, such as asthma, bronchitis, and allergic conditions, have been reported to have a lower prevalence in samples of care-experienced children [20, 21]. However, the evidence for this lower prevalence is mixed for these and other conditions [13, 16]. A further complication is that care-experienced children come disproportionately from families exposed to greater socioeconomic disadvantage [17]. This independently contributes to poor health and should ideally be accounted for when studying the health outcomes of these children. Clarifying our understanding of the physical health problems which disproportionately affect young care-experienced children is an important first step towards decreasing the inequalities these children are subjected to.

To the best of our knowledge, the literature regarding the physical health outcomes of young care-experienced children in high-income countries has not been reviewed. However, there have been several reviews regarding other groups of care-experienced children and other aspects of the health of these groups. Lee et al. carried out a systematic review of physical health outcomes of foster children which focussed on ethnic and racial inequalities [22]. Dental health outcomes and behaviours have also been the subject of multiple recent reviews [19, 23, 24]. Other reviews have looked at developmental and mental health [25–28], as well as engagement with health care services [29–31]. Most of these works (and many studies cited above) combine care-experienced children of all ages. However, the type of health conditions an individual experiences varies substantially between infancy, middle childhood, and adolescence. Therefore, there is a need to review the research relating specifically to physical health in the early years.

An important caveat regarding differences in health between care-experienced children and those without experience of care are cases where serious health conditions are the primary reason for children being in care. Epilepsy and cerebral palsy, for example, have been found to be substantially more common in children in care [16, 21]. Severe forms of these conditions can require intensive management which cannot reasonably be provided in a typical family home. However, the substantial majority of children enter care due to being subjected to parental abuse or neglect [32–35]. Children entering care primarily due to complex health conditions have, therefore, had socioenvironmental experiences which are not representative of most children in care. In turn, the type of health conditions that affect children in care for medical reasons are likely to be substantially different to the majority of children in care. As such, it is young children that are, or were, in care for non-medical reasons which our work is concerned with.

### 1.1 Study aims

The aim of this work was to collate, review, and synthesise the results of peer-reviewed studies which examined the physical health outcomes of young care-experienced children. The review addressed the question: “Which physical health conditions have been studied or observed to affect young care-experienced children in high-income countries?” and subsequently, where possible: “How do these health conditions affect children in care compared to children without care experience?”. By synthesising the results of this literature we aimed to provide a resource that would be useful to policymakers, healthcare professionals, as well as providing context and recommendations for future research.

## 2 Materials and methods

The scoping review method used here provides a systematic way of combining heterogeneous sources of evidence [36–39]. We selected this method as we expected a broad range of physical health outcomes and it can accommodate international heterogeneity in the definition of care. To outline the purpose of the review and the inclusion criteria we first generated a Population Concept Context (PCC; Table 1) framework [40]. The five-stage method of Arksey et al. [36], the refinements of Levac et al. [41], and Joanne Briggs Institute guidance [38] informed the design of the review. Further details are detailed in a previously published protocol paper [42]. Our work also follows the *Preferred Reporting Items for Systematic Reviews and Meta-Analyses: Extension for Scoping Reviews* guidelines [43], the checklist for which is provided in S1 Table.

**Table 1:** Details of the Population, Concept, and Context framework.

|  |  |
| --- | --- |
| Population | Care-experienced children who meet all following criteria:<br>- Aged less than seven years<br>- Not entering care during the first three months of life<sup>a</sup><br>- Not in care primarily for health-related reasons<br>- Have experience of any of the following care settings:<br>- Parental care with additional social work input<br>- Kinship care<br>- Foster care<br>- Residential care<br>- Adoption or prospective adoption |
| Concept | Health outcomes primarily expressed or rooted in physiology, including mortality |
| Context | High-income countries<sup>b</sup> |

Notes: <sup>a</sup>The health issues affecting children that enter care neonatally are an important subject for research. However, we chose to exclude studies where the sample was comprised solely of children entering care at this age since the health conditions to which they are susceptible are mainly reflective of prenatal conditions. <sup>b</sup>Children living in high-income countries face challenges to their health which are substantially and qualitatively different than children in low- and middle-income countries [44,45]. Particularly, the burden of health problems in high-income countries skews towards non-communicable diseases.

### 2.1 Inclusion criteria

Studies were included if the participants were currently living in or had previously lived in one of the following care settings: parental care while receiving additional formal support from social services, kinship care, foster care, residential care, or having been adopted. Studies were included if the children were living in high-income countries (as per The World Bank national categorisation [46]). Children were required to be under seven years of age and from samples not predominantly comprised of neonatal adoptees. The latter are largely affected by prenatal conditions rather than early childhood experiences. Studies of children outside this age range were included where results were provided for subgroups within this range. Health outcomes were restricted to the following: health conditions assessed or diagnosed by a health professional; parental report of a current or previous diagnosis by a health professional; assessment by validated scales, diagnostic manuals, or health assessment surveys. Studies using bespoke scales relating to physical health were included at the discretion of reviewers. Studies must have been published in peer-reviewed journals as original research items and indexed by the selected databases prior to 1st October 2024.

### 2.2 Exclusion criteria

Studies where the sample was completely or majority comprised of children that entered care primarily due to complex physical health needs were excluded to avoid confounding. Studies reporting disease outbreaks in residential care accommodation were also excluded since the physical health of children living there was incidental and not informative for the present review. Studies were also excluded where the sample was completely or majority comprised of children that entered care at birth (e.g., neonatal adoptees) as their health is more reflective of maternal health and the prenatal environment. Studies whose only outcomes were physiological markers not directly associated with a disease were also excluded (e.g., reports of salivary cortisol levels were excluded, but reports of low serum ferritin were included as this is a marker of anaemia).

Studies where most or all of the children in the sample were adopted from non-high-income countries were excluded. This was due to qualitative differences in health problems experienced by children in non-high-income countries compared to children in high-income countries [47]. Articles that did not have English language titles and abstracts were excluded for pragmatic reasons.

### 2.3 Searches

Keywords relating to elements of the PCC framework described in Table 1 were used to create a preliminary search strategy. Additional keywords identified in the results of pilot searches on the MEDLINE database were added until saturation was reached.

Final strategies were developed for the MEDLINE, CINAHL, and Web of Science Core Collection databases and applied to items indexed before 1st October 2024. These search strategies were developed with the assistance of a specialist information scientist at the Medical Research Council/Chief Scientist’s Office Social & Public Health Sciences Unit at the University of Glasgow. Complete search strategies for all databases are included in the supporting information (S2 Table). Reference lists of key papers known to the authors were also searched manually for relevant titles. Duplicate search results were removed using Microsoft EndNote and Rayyan [48]. Full texts were gathered via online journal repositories, Google Scholar, ResearchGate, direct requests to authors, searches of institutional repositories, and inter-library loans.

### 2.4 Screening, data extraction, and quality assessment

Title and abstract screening were completed concurrently in Rayyan [48]. Duplicates were removed using deduplication functions and manual confirmation. One reviewer (DRRB) screened all items and a second reviewer (AS) independently screened a 20% random sample after an initial training set were screened collaboratively. Two reviewers (DRRB and AS) screened all available full-text items except for non-English-language papers. Non-English items were translated and screened by native speakers, with machine-translated versions also screened by DRRB where possible. In cases of ambiguity at any screening stage the item was passed forward to the next stage for more comprehensive assessment. Disagreements were resolved by discussion.

Data from included full-text items were recorded on a bespoke extraction form (see S3 Table). Bibliographic data, study methods, and health outcomes were tabulated to identify patterns and clusters in each of these domains. The collated results were reported using a narrative synthesis. Numerical data in the included studies were reported with varying levels of precision. Proportions were calculated to one decimal place and reported here when count data were available from the included study. If proportions were originally reported to the nearest whole percentage without count data, this level of precision was retained when reporting the result(s) here. In some cases, proportions were reported along with the total sample size but without count data for the specific health outcome. In other cases, proportions were provided alongside the number of children affected by a specific outcome but without a sample size. In these cases either the approximate number of children with the specified outcome or the sample size was estimated. Count data or sample sizes which have been estimated in this way are reported below using an ‘approximately equal to’ symbol (≈).

A brief, bespoke quality assessment checklist was developed based on variables from the data extraction form. Its purpose was to highlight high-quality studies that may be of particular interest to readers. High-quality studies were defined as those meeting the following criteria:

- Inclusion of a comparator group
- Analyses adjusted for age, sex, and socioeconomic status (SES) or the comparator group was selected to be of similar demographic constitution (e.g., children living in areas of lower SES receiving subsidised or social healthcare in the USA).
- Physical health was a primary focus of the study
- Sample size of at least 30 care-experienced children and 30 comparator children

## 3 Results

Database searches identified 17,363 unique items and seven additional items were identified by manual searches of citation titles in review papers that were known to the authors [33, 49–54]. After screening titles and abstracts, 179 items were included for full-text screening, of which 164 were available. After full-text screening, 36 relevant items were included in the review [12, 17, 18, 33, 49, 51, 52, 55–83]. Details of the screening process are provided in the PRISMA flowchart in Fig 1.

**Fig 1.** PRISMA-ScR flowchart.

### 3.1 Study characteristics, participants, and settings

A summary of the basic details of the 36 studies that were included and participant demographics are presented in Table 2 and Fig 2. Over three-quarters of studies included mixed care settings or did not report the specific type of care setting (n = 29, 80.6%); five studies (13.9%) were carried out in residential care settings; one study (2.8%) considered adopted children; one study (2.8%) considered children in kinship care. Notably, no studies were identified that examined children in care while remaining in the parental home, and very few studies considered children in kinship care. This may be attributable to the substantial number of studies reporting mixed care types and to the broad, often ambiguous, definition of foster care in the USA, where the majority of studies were undertaken. The USA was the most common setting (n = 21, 58.3%) followed by the UK (n = 5, 13.9%) and Australia (n = 2, 5.6%). One study (2.8%) was identified from each of Canada, France, Israel, Portugal, Russia, Saudi Arabia, Spain, and Sweden. The studies were published between 1955 and 2024: 11 before 2000 (30.6%); eight in 2000–2009 (22.2%); and 17 in 2010 or later (47.2%). Six studies had a longitudinal component (16.7%) but the majority were cross-sectional (n = 30, 83.3%). Studies were published in 26 different journals, with *Pediatrics* being the most common source (n = 4, 11.1%).

**Fig 2.** Distribution of included studies. Distribution of included studies by (A) decade of publication, (B) country, and (C) care setting. Studies were categorised into a single care setting. *Foster (USA)* denotes studies conducted in the United States where “foster care” was used as a general term without specifying the type of placement, which may include kinship, residential, or non-relative foster care. *Mixed* refers to studies that explicitly included more than one care setting (e.g., both foster and residential care) but did not disaggregate results by setting. *Not reported* indicates that no specific care setting was described.

**Table 2:** Characteristics of included studies and their samples.

| Study | Data period | Country | Care type | Comparator group | Sample size | % female | Age (mean) | Age (min.) | Age (max.) |
| --- | --- | --- | --- | --- | --- | --- | --- | --- | --- |
| AlSadhan et al. (2017) [55] | - | Saudi Arabia | Residential | Y | 25 | - | - | 48 | 83 |
| Arcot et al. (2023) [56] | 2010–2015 | USA | Foster<sup>a</sup> | N | 28,930 | - | - | 6 | 59 |
| Bel et al. (2002) [57] | 1995–1998 | Spain | Residential | N | 82 | - | - | 0 | 107<sup>b</sup> |
| Bramlett et al. (2007) [18] | 2003 | USA | Adopted | N | - | - | - | 0 | 71 |
| Calheiros et al. (2022) [58] | - | Portugal | Residential | N | 100 | 51 | - | 0 | 83 |
| Chaiyachati et al. (2020) [59] | 2003–2016 | USA | Foster<sup>a</sup> | Y | 2,269,561PY<sup>c</sup> | - | - | 12 | 59 |
| Chernoff et al. (1994) [49] | 1989–1991 | USA | Foster<sup>a</sup> | N | 408 | - | - | 0 | 35 |
| Croft et al. (2011) [60] | 2008 | UK | -<sup>d</sup> | N | 15 | - | - | 25 | 59 |
| de Montaigne et al. (2015) [61] | - | France | Mixed | N | 40 | 35 | - | 0 | 83 |
| Dhindsa et al. (2020) [62] | 2015–2018 | UK | -<sup>d</sup> | Y | 9 | - | - | 24 | 59 |
| Dubowitz et al. (1992) [63] | 1989 | USA | Kinship | N | 137 | - | - | 0 | 71 |
| Ferguson et al. (2006) [64] | 2000 | USA | Foster<sup>a</sup> | N | 92 | - | - | 24 | 71 |
| Greiner et al. (2017) [65] | 2012–2015 | USA | Foster<sup>a</sup> | N | 640 | - | - | 6 | 83 |
| Halfon et al. (1995) [66] | 1988–1991 | USA | Foster<sup>a</sup> | N | 185 | - | - | 0 | 60 |
| Hansen et al. (2004) [51] | 1998–1999 | USA | Foster<sup>a</sup> | N | 80 | - | - | 0 | 47 |
| Hill (2003) [67] | 2001 | UK | -<sup>d</sup> | N | - | - | - | 0 | 59 |
| Högberg et al. (2019) [12] | 1997–2014 | Sweden | -<sup>d</sup> | N | 1,514 | - | - | 0 | 12 |
| Lanais et al. (2024) [68] | 2009–2021 | Australia | Mixed | Y | 666 | - | - | -<sup>e</sup> | -<sup>e</sup> |
| Lee et al. (2024) [69] | 2009–2019 | USA | Mixed | N | -<sup>f</sup> | - | - | 0 | 59 |
| Leggat et al. (2024) [70] | 2011–2018 | Australia | Mixed | N | 1,082 | - | - | 9 | 66 |
| Leslie et al. (2005) [33] | 1998–1999 | USA | Foster<sup>a</sup> | N | 1,542 | 47 | 35.2 | 3 | 71 |
| McLeigh et al. (2023) [71] | 2017–2020 | USA | Mixed | N | 1,072 | 45.2 | - | 36 | 71 |
| McMahon et al. (2018) [17] | 2007–2013 | UK | -<sup>d</sup> | Y | 894 | - | - | 60 | 71 |
| Miller et al. (2006) [72] | - | Russia | Residential | N | 127 | - | 21.3 | 1.5 | 72 |
| Payne et al. (1998) [73] | - | UK | -<sup>d</sup> | Y | 62 | - | - | 24 | 71 |
| Radel et al. (2023) [74] | 2019 | USA | Mixed | Y | 124,351 | - | - | 36 | 71 |
| Schneiderman et al. (2013) [75] | 2006–2010 | USA | Mixed | N | 83 | - | - | 24 | 71 |

Table 2: Characteristics of included studies and their samples. (Continued)
| Study | Data period | Country | Care type | Comparator group | Sample size | % female | Age (mean) | Age (min.) | Age (max.) |
| --- | --- | --- | --- | --- | --- | --- | --- | --- | --- |
| Siefert et al. (1994) [76] | 1980–1989 | USA | Foster<sup>a</sup> | N | - | - | - | 1 | 12 |
| Simms (1989) [77] | 1985–1987 | USA | Foster<sup>a</sup> | N | 113 | 44.2 | 31.2 | 1 | 72 |
| Swire et al. (1977) [78] | 1973–1974 | USA | Foster<sup>a</sup> | N | - | - | - | - | 71 |
| Swire et al. (1978) [79] | 1971–1974 | USA | Foster<sup>a</sup> | N | 307 | - | - | - | 71 |
| Takayama et al. (1998) [80] | 1991–1992 | USA | Foster<sup>a</sup> | N | 361 | - | - | 0 | 83 |
| Tenenbaum et al. (2011) [81] | - | Israel | Mixed | N | 100 | 58 | 5.9 | 0.5 | 24 |
| Valencia-Rojas et al. (2008) [52] | 1991–2004 | Canada | Residential | Y | 66 | 43.9 | 49.2 | 24 | 83 |
| Wallace et al. (1955) [82] | - | USA | Foster<sup>a</sup> | N | 210 | - | - | 0 | 71 |
| Wyatt (1997) [83] | 1992–1994 | USA | Foster<sup>a</sup> | N | 45 | - | 35 | 18 | 83 |

*Notes:* Blank cells indicate no data for this attribute were reported. *Sample size* reports the number of care-experienced children in the study. *Age* values reported in months. <sup>a</sup>The term *foster care* in the USA incorporates all forms of care described in Table 1 with the exception of adoption. <sup>b</sup>Bel et al. [57] included children aged 0–8 years. The reviewers felt the results were relevant to the research question and the study was of good quality despite some children not meeting the age inclusion criteria. <sup>c</sup>PY = person-years. <sup>d</sup>These studies described the sample as being composed of children in care without describing the distribution of children in different care settings. <sup>e</sup>Children in this sample were predominantly aged 5.4–6.2 years (73.7%) and the remainder were slightly younger or older but within the same school year cohort. <sup>f</sup>All subgroup results for younger children were presented as relative rates compared to the reference category of children aged 15–17 years.

### 3.2 Study outcomes

The physical health of care-experienced children was a primary outcome in 35 (97.2%) studies and a secondary outcome in one study (2.8%). In the study where health was a secondary outcome a single-item assessment of general health was included as part of the primary objective of analysing socioenvironmental risk factors for entering care [58]. Many of the included studies focused on specific and limited sets of health outcomes (69.4%, n = 25) with most other studies reporting on broader multi-system health reviews (22.2%, n = 8). Assessment by clinical professionals and, separately, reviewing medical records were the most common methods reported (both 38.9%, n = 14), followed by objective measurement of height and/or weight (22.2%, n = 8). A summary of the results of the included studies is provided in Table 3 and Fig 3. See S3 Table for more details on the distribution of health outcomes and the methods used in the included studies. Note that some studies used multiple methods. so percentages may sum to greater than 100%.

**Fig 3.** Number of studies reporting specific health outcomes among children in care. Subcategories within broader outcome groups (e.g., *Weight - Obesity* ) are indicated by a hyphen, a lighter shade and with italicised labels. Subcategories are positioned directly below their respective parent categories.

**Table 3:** Summary of key outcomes reported in included studies.

| Study | Health outcomes | Study methods | Summary of main findings |
| --- | --- | --- | --- |
| AlSadhan et al. (2017) [55] | Dental health | Clinical assessment | Higher prevalence of tooth trauma (predominantly tooth fracture) in children in a comparator group (28.0%, n = 7 of 25) than children in care (16.0%, n = 4 of 25) but the small sample size limits generalisability. |
| Arcot et al. (2023) [56] | Haematological | Medical records review | In children aged six months to one year, 3.1% (n = 411 of 13,323) had a diagnosis of anaemia. In children aged 2–4 years, this increased to 3.9% (n = 610 of 15,607). |
| Bel et al. (2002) [57] | Height; weight; head circumference | Objective height/weight measurement | Many of the sample of children entering care aged under two years were more than two standard deviations below expected mean value for height (45.5%, n = 20 of 44), weight (20.5%, n = 9 of 44), and head circumference (22.7%, n = 10 of 44). These proportions reduced significantly by the time these children exited care to 31.8% for height (n = 14 of 44), 13.6% for weight (n = 6 of 44) and, 13.6% for head circumference (n = 6 of 44), respectively. Poor physical development (more than two standard deviations below expected mean value) was less common at entry to care for children aged 2–8 years (height: 13.2%, n = 5 of 38; weight: 2.6%, n = 1 of 38; head circumference not measured beyond infancy) with necessarily smaller reductions in proportion by the time children exited care (height: 10.5%, n = 4 of 38; weight: 0.0%, n = 0 of 38.) |
| Bramlett et al. (2007) [18] | Accident and injury | Parental report | After adjusting for significantly different demographic factors, adopted children aged 0–5 were less likely to have sustained an injury requiring medical treatment in the previous year than a comparator group without care experience - 7.4% compared to 9.8%. N.B. These proportions arise from multivariable models adjusted for relevant covariates so it is not appropriate to report count data here. |
| Calheiros et al. (2022) [58] | General health | Clinical assessment | Study objective was to assess socioenvironmental risk factors for children entering care and build profiles of emergent clusters. Of the 100 children assessed, 35.0% (n = 35) were deemed to have poor general physical health. |
| Chaiyachati et al. (2020) [59] | Mortality | Administrative data | Adjusted mortality rates in children aged 1–4 years in the USA were 50.7 per 100,000 person-years (95% CI: 47.8, 53.6) for children in care and 27.5 per 100,000 person-years (95% CI: 27.3, 27.7) for children not in care; a rate ratio of 1.84 (95% CI: 1.74, 1.95). |
| Chernoff et al. (1994) [49] | Head circumference; height; immunisation; weight | Clinical assessment; medical records review; objective height/weight measurement | In the children aged under three years, disproportionately high numbers of children were in or below the 5th percentile for height (26.2%, n ≈ 107 of 393), weight (15.4%, n ≈ 61 of 396), and head circumference (18.8%, n ≈ 73 of 388). Of children under six years, 30.8% (n = 194 of 630) were insufficiently immunised for age. N.B. Sample sizes varied for each measurement due to children being "... too young, ... uncooperative ... or because of time constraints." |
| Croft et al. (2011) [60] | Obesity; weight | Medical records review; objective height/weight measurement | Small sample of children in care aged 2–5 years, of which 9.4% (n ≈ 3 of 32) were considered to be overweight or obese. |

Table 3: Summary of key outcomes reported in included studies. (Continued)
| Study | Health outcomes | Study methods | Summary of main findings |
| --- | --- | --- | --- |
| de Montaigne et al. (2015) [61] | Gastrointestinal; hearing; motor control; vision | Database review | The distribution of 1,063 health problems across 484 children in care (of all ages) was reported. However, the total number of health problems among the subsample of 40 children aged six or under was not specified. Consequently, the following proportions cannot be used to determine the number of children in this age group affected by each condition. Among children aged six or under, health problems were distributed as follows: 6.7% had non-specific gastrointestinal issues, 2.7% had hearing problems, 2.8% had motor control problems and, 10.7% had visual impairments. The remaining 79.8% of diagnoses in this group were primarily related to psychosocial issues. |
| Dhindsa et al. (2020) [62] | Obesity; weight | Objective height/weight measurement | Small sample of children in care aged 2–4 years. Two children of nine (22.2%) were classed as overweight using clinical centiles, three (33.3%) using population centiles. |
| Dubowitz et al. (1992) [63] | Cardiovascular; dental health; dermatological; failure to thrive; haematological; hearing; height; immunisation; obesity; respiratory; vision; weight | Clinical assessment; health survey; medical records review | Failure to thrive was identified in 9% ($n \approx 12$ of 137) children in kinship care. Absolute values for height showed a disproportionately high number of children in the 5th percentile or below (9%, $n \approx 12$ of 137). Obesity, defined as weight-for-height in 96th percentile or above, was noted in 8% ($n \approx 11$ of 137) children.<br>Dental caries reported in 15% of children ($n \approx 21$ of 137) and 7% ($n \approx 10$ of 137) reported to have “other dental” problems. Children may have received one or either of these diagnoses.<br>Atopic dermatitis specifically was identified in 5% ($n \approx 7$ of 137) of children. More broadly, 15% ($n \approx 21$ of 137) of children had any dermatological issue.<br>Vision and hearing were tested in 55 and 65 children aged 3–5 years and failure rates were reported at 16% ($n \approx 9$ of 55) and 6% ($n \approx 4$ of 65), respectively. However, a substantial proportion of the children in the wider sample children were classified as untestable due to requiring vision or hearing aids but not having them at the time of screening.<br>Asthma was identified in 26% ($n \approx 36$ of 137) of children.<br>Appropriate immunisation was observed in 56% ($n \approx 77$ of 137) of children but nearly all of the remaining children were conservatively classed as having an uncertain immunisation record (as opposed to being known to be incompletely immunised).<br>Anaemia was treated or suspected in 9% ($n \approx 12$ of 137) of children tested. A small fraction of children assessed were noted to have high blood pressure (4%, $n \approx 3$ of 77 children assessed). |

Table 3: Summary of key outcomes reported in included studies. (Continued)
| Study | Health outcomes | Study methods | Summary of main findings |
| --- | --- | --- | --- |
| Ferguson et al. (2006) [64] | Cardiovascular; medication; neurological | Medical records review | Study examined prescriptions for a range of psychotropic medications (and medications treating side effects of psychotropics). Identified only one child in a sample of 92 (1.1%) aged 0–6 years who received a prescription of anti-hypertensive medication to treat migraines. |
| Greiner et al. (2017) [65] | Haematological; infectious disease | Laboratory testing of samples | High concentrations of lead were found in the blood of 2.7% ($n \approx 17$ of 640) of children in care aged six months to six years. However, the authors noted the laboratory equipment used to assess these concentrations was later recalled due to calibration error resulting in falsely-low measurements. This would suggest the reported prevalence is an underestimate.<br>No cases of tuberculosis were detected following skin tests. |
| Halfon et al. (1995) [66] | Abuse; accident and injury; allergies; burns; dermatological; failure to thrive; gastrointestinal; genitourinary; haematological; hearing; height; infectious disease; motor control; neurological; respiratory; vision; weight | Medical records review | The following prevalences of health conditions were reported in the sample of 185 children aged under 61 months entering care: 22.2% ($n = 41$) had anaemia; 4.9% ($n = 9$) had a non-specific cardiovascular disorder; 21.6% ($n = 40$) had a dermatological condition; 17.8% ($n = 33$) were diagnosed with asthma; 13.5% ($n = 25$) had a gastrointestinal disorder; and injuries requiring medical attention were found in 1.1% ($n = 2$) of children.<br>Disproportionately high numbers of children aged 0–4 years exhibited poor physical development: 25.4% ($n = 47$) under 5th percentile for height, 36.2% ($n = 67$) under 25th percentile; 16.8% ($n = 31$) under 5th percentile for weight; 11.9% ($n = 22$) had head circumferences under 5th percentile; and 10.3% ($n = 19$) were labelled as failing to thrive.<br>Gross motor control problems were identified in 34.6% of children ($n = 64$) and fine motor control problems in 36.2% ($n = 67$).<br>(Only a subset of results are summarised here due to the large number of outcomes reported in this paper. See the dedicated worksheet in S1 Table for more information on the outcomes of this paper.) |
| Hansen et al. (2004) [51] | Head circumference | Medical records review | Slightly fewer than expected children had head circumferences at or below the 25th percentile (16.3%, $n = 13$ of 80). |
| Hill (2003) [67] | Immunisation | Administrative data; database review | Meningococcal C immunisation rates were lower in children in care aged 0–4 years when aggregated across several regional health boards (risk ratio = 1.79). There was substantial variation between boards and in some cases children in care had higher rates of immunisation (range of risk ratios from 0.53 to 2.48). However, this was a convenience sample composed of a selection of non-representative geographic districts. |

Table 3: Summary of key outcomes reported in included studies. (Continued)
| Study | Health outcomes | Study methods | Summary of main findings |
| --- | --- | --- | --- |
| Högberg et al. (2019) [12] | Abuse; accident and injury; bone fracture; burns; failure to thrive; neurological | Administrative data | In a sample of children in care that had received inpatient treatment for outcomes suspected to relate to abuse, 41.9% had been treated for head injury and/or craniocerebral conditions, representing 4.4% of care-experienced children in the sample (n = 328 of 7,455). Bone fractures occurred in 16.9% of the inpatient cases but only 1.8% of the full sample (n = 131 of 7,455). Failure to thrive was diagnosed in 9.0% of the inpatient cases but only 0.9% of the full sample (n = 70 of 7,455). Seizures were diagnosed in 6.8% of the inpatient cases but only 0.9% of the full sample (n = 70 of 7,455). Burns were rare and observed in only 0.1% of all children (n = 6 of 7,455). |
| Lanais et al. (2024) [68] | General health | Clinical assessment; health survey | Using a standardised health and development assessment tool, 38% (n ≈ 127 of 333) male children and 27% (n ≈ 90 of 333) of female children that had entered out-of-home-care were classified as being “vulnerable” in the physical health domain (indicating poor physical health). By comparison, only 14% and 6% (sample sizes not reported) of male and female children, respectively, that had never been involved with child protective services were classed as “vulnerable”. Note: no explicit definition of “vulnerable” was provided but this classification appears to derive from normative general population data. |
| Lee et al. (2024) [69] | Mortality | Administrative data | Children in care aged 1–4 years were not at any increased risk of mortality compared to the reference group (children in care aged 15–17 years; incident rate ratio = 1.00 [95% CI: 0.96, 1.04]). Children in care aged under one year were at increased risk (incidence rate ratio = 1.06 [95% CI: 1.02, 1.11]) but it is unclear if this age category included deaths of newborn children removed from parents at birth. No comparisons were made with children in the general population. |
| Leggat et al. (2024) [70] | Motor control | Clinical assessment | Across the whole sample, including children with and without a history of exposure to parental substance use, 20.0% (n ≈ 217 of 1,082) of children had atypical gross motor development and 25.9% (n ≈ 280 of 1,082) had atypical fine motor development. Results were broken down by history versus no history of parental substance use. Where parental substance use had been present, 15.6% of children (n ≈ 89 of 563) had gross motor problems and 22.0% (n ≈ 124 of 563) had fine motor problems. Where parental substance use was not present, rates were higher with 24.7% of children (n ≈ 128 of 519) having gross motor problems and 30.1% (n ≈ 156 of 519) having fine motor problems. |
| Leslie et al. (2005) [33] | Abuse; cardiovascular; dental health; dermatological; gastrointestinal; genitourinary; haematological; hearing; musculoskeletal; neurological; respiratory; vision | Clinical assessment; medical records review | Nearly all children (86.7%, n = 1,337 of 1,542) had a physical health condition identified at a health screening upon entry to care. Prevalence of conditions by organ system: dermatological - 66.5% (n = 1,026 of 1,542), respiratory - 22.6% (n = 348 of 1,542), dental - 13.2% (n = 204 of 1,542), anaemia - 7.4% (n = 114 of 1,542), auditory - 6.7% (n = 104 of 1,542), substantiated non-accidental trauma - 6.2% (n = 96 of 1,542), vision - 3.3% (n = 51 of 1,542), musculoskeletal - 2.9% (n = 44 of 1,542), neurological - 2.5% (n = 38 of 1,542), cardiovascular - 2.3% (n = 36 of 1,542), genitourinary - 1.9% (n = 29 of 1,542), gastrointestinal - 1.6% (n = 24 of 1,542). |

Table 3: Summary of key outcomes reported in included studies. (Continued)
| Study | Health outcomes | Study methods | Summary of main findings |
| --- | --- | --- | --- |
| McLeigh et al. (2023) [71] | General health | Medical records review | In a review of medical records of children aged 3–5 years that had attended one of two primary care clinics for foster children in the USA, 36.8% of children (n = 394 of 1,072) had non-complex but chronic physical health status. A further 22.4% (n = 240 of 1,072) had complex chronic physical health status. The remainder (40.9%, n = 438 of 1,072) were classified as having non-complex, non-chronic physical health status. |
| McMahon et al. (2018) [17] | Dental health | Administrative data | Just under one quarter of looked after children (23.4%, n = 209 of 894) undergoing a universal dental review at age five years required urgent dental treatment. Adjusted odds ratio comparing children in care to those not in care was 2.06 (95% CI: 1.76, 2.42). Considering both urgent and non-urgent dental needs two-thirds of looked after children (66.6%, n = 595 of 894) required further treatment, yielding an adjusted odds ratio of 2.65 (95% CI: 2.30, 3.05) compared to children not in care. |
| Miller et al. (2006) [72] | Cardiovascular; cleft palate; Down's syndrome; foetal alcohol syndrome; head circumference; height; vision; weight | Clinical assessment; objective height/weight measurement | Foetal alcohol syndrome was suspected or confirmed in 48.0% of children (n ≈ 96 of 201 screened) in a residential care unit, 6.4% (n = 15 of 234) had Down's syndrome, and 2.1% (n = 5 of 234) had congenital heart disease.<br>Physical development in children was severely restricted with mean z-scores of -1.40, -1.59, and -1.38 for height, weight, and head circumference, respectively. |
| Payne et al. (1998) [73] | Immunisation | Database review; medical records review | Children in care had lower rates of immunisation than an unmatched comparator group of children not in care, although around one-third of the care-experienced sample were missing immunisation records. Immunisation rates: diphtheria, tetanus, polio (primary) – 90.3% (n = 56 of 62) compared to 96.9% (n = 23,197 of 23,919); diphtheria, tetanus, polio (booster) – 32.2% (n = 20 of 62) compared to 43.4% (n = 10,383 of 23,919); pertussis – 67.7% (n = 42 of 62) compared to 89.8% (n = 21,482 of 23,919); measles, mumps, and rubella – 85.4% (n = 52 of 62) compared to 94.7% (22,672 of 23,919); and haemophilus influenzae type b – 46.7% (n = 29 of 62) compared to 70.7% (n = 16,907 of 23,919). |
| Radel et al. (2023) [74] | Medication | Administrative data; database review | In children in care aged 3–5 years, ~6.5% were prescribed a psychotropic medication compared to ~3.0% of children not in care but enrolled in Medicaid (i.e., living in comparable socioeconomic environments). The proportion of children receiving medications varied by class (from samples of 124,351 children in care and 6,650,494 children not in care): antipsychotics – 1.06% versus 0.45%; anticonvulsants – 0.65% versus 0.22%; antiparkinsonian – 0.02% versus <0.01%; CNS 0.05% versus 0.01%. |
| Schneiderman et al. (2013) [75] | Height; obesity; weight | Objective height/weight measurement | Obesity (weight in or above the 95th percentile) and overweight (weight in or between the 85th and 94th percentiles) were observed in 14.5% (n = 12 of 83) and 13.3% (n = 11 of 83), respectively, of a sample of children aged 2–5 years. |

Table 3: Summary of key outcomes reported in included studies. (Continued)
| Study | Health outcomes | Study methods | Summary of main findings |
| --- | --- | --- | --- |
| Siefert et al. (1994) [76] | Mortality | Database review | Statewide standardised mortality rates averaged over the period 1980–1989 were almost double for children in their first care placement compared to those not in care (rate ratio = 1.90). Deaths were yet more frequent among “African American” children compared to the statewide non-care rate (rate ratio = 2.46). |
| Simms (1989) [77] | General health; height; motor control | Clinical assessment; objective height/weight measurement | Chronic health problems were observed in 35.4% (n = 40 of 113) of children. Acute health conditions were less commonly identified (10.6%, n = 12 of 113).<br>A disproportionately high number of children had heights in the lowest five percentiles (14.2%, n = 16 of 113).<br>Gross motor control problems were observed in 23.9% (n = 27 of 113) of children. Fine motor control problems were observed in 29.2% (n = 33 of 113) children. |
| Swire et al. (1977) [78] | Dental health; general health; height; weight | Clinical assessment; medical records review | A mean of 0.76 chronic physical health conditions per child aged under six years was identified during health screening, a higher frequency than those aged over six.<br>More children were in the lowest five percentiles for height and weight than expected (11%, age subgroup sample size was not reported).<br>Dental treatment was required by 21% of the children aged 3–5 years and decay (with or without need for treatment) was reported in “one-third” of this age group. |
| Swire et al. (1978) [79] | Immunisation | Clinical assessment; medical records review | Immunisation status was unknown or behind expected schedule for mumps in 68% of “preschool” children, 36% for measles, 43% for rubella, 19% for poliomyelitis, and 23% for diphtheria, tetanus and pertussis. N.B. The sample size of “preschool” children was not reported but around 307 children from the overall sample were under 6 years. |
| Takayama et al. (1998) [80] | Abuse; accident and injury; dental health; dermatological; haematological; hearing; infectious disease; neurological; respiratory; vision | Laboratory testing of samples; medical records review | Prevalence estimates derived from broad health reviews carried out within 72h of a child entering care: recent injury - 5.8% (n ≈ 31 of 361), “marks of abuse” - 4.4% (n ≈ 16 of 361), dermatological conditions - 20.5% (n ≈ 74 of 361), anaemia - 9.9% (sample size reported as ≤ 309), dental caries - 4.0% (sample size reported as ≤ 309), upper respiratory illness - 27.4% (n ≈ 99 of 361), asthma - 3.6% (n ≈ 13 of 361), neurological conditions - 2.2% (n ≈ 8 of 361).<br>No hearing problems were identified in the subgroup of children aged 3–6 years and 8.6% of these children had abnormal vision (tested without vision aids) but the sample size was ambiguously reported as ≤ 309 for this age subgroup. |

Table 3: Summary of key outcomes reported in included studies. (Continued)
| Study | Health outcomes | Study methods | Summary of main findings |
| --- | --- | --- | --- |
| Tenenbaum et al. (2011) [81] | Failure to thrive; foetal alcohol syndrome | Clinical assessment; laboratory testing of samples | Facial and cranial morphology consistent with foetal alcohol syndrome was observed or prospectively diagnosed in 4.0–6.0% of children aged under two years referred to a health clinic for children entering care. The variation in proportion is due to the subjective nature of foetal alcohol syndrome diagnosis. Failure to thrive was also noted in 15.0% (n = 15 of 100) of the sample. |
| Valencia-Rojas et al. (2008) [52] | Dental health | Medical records review | The majority of children in the sample had signs of dental caries (57.6%, n = 38 of 66) and severe forms of caries were also common (31.8%, n = 21 of 66). The prevalence of dental caries was approximately double that found in a comparable group of children not in care in a separate study (57.6% compared to 30%, sample size not reported for the latter). All children in the care-experienced sample had been selected as they were subject to abuse or neglect. Further, the comparison group was not matched for gender or SES but was from the same city. Dental trauma was relatively rare with only 6.1% of children (n = 4 of 66) displaying symptoms. |
| Wallace et al. (1955) [82] | Vision | Clinical assessment | In children aged under two years, 24.2% (n = 15 of 62) had a vision problem with the most common issue being refractive errors, particularly hyperopia. In children aged 2–6 years, 29.7% (n = 44 of 148) had a vision problem with refractive errors again being the most common problem. |
| Wyatt (1997) [83] | Height; weight | Clinical assessment; objective height/weight measurement | Upon entry to care, the sample of 62 children were small for weight and height with mean z-scores of -0.16 and -0.21, respectively. Children were followed-up after one year, regardless of whether they returned to the parental home, with a mean time in care of 9.5 months. Both growth measures improved significantly by follow-up. By contrast, weight-for-height was relatively high at both time points (z = 0.30 and 0.40). |

*Notes:* <sup>a</sup>Percentages extracted from the included studies are reported throughout to one decimal place except in cases where the original studies rounded to whole numbers and underlying counts were not available. This means some figures may have a trailing zero but others may not.

### 3.3 Physical health

Studies which carried out broad health reviews reported a wide range of estimates for the proportion of children experiencing any physical health issue at the point of review, varying from 35% to over 85% [33, 58, 66, 77], highlighting the large heterogeneity across studies. When classifying children in care’s complexity and chronicity of physical health issues, McLeigh et al. observed 22.4% (n = 240 of 1,072) of children had complex chronic health issues and a further 36.8% had non-complex chronic health issues [71]. Swire et al. reported a mean of 0.76 chronic problems per child which were expected to have enduring effects on health [78]. However, Halfon et al. observed that the number of distinct health conditions per child decreased with age and reported a mean of 2.6 conditions aged under one year, 1.5 problems aged 1–3 years, and 1.3 problems for children aged 3–5 years [66]. Lanais et al. reported that 38% of male children (n ≈ 127 of 333) and 27% of female children (n ≈ 90 of 333) in out-of-home care were considered “vulnerable” on the physical health domain of a standardised development assessment tool, compared to just 14% and 6% of children with no history of involvement with care or child protective services (sample size not reported; no definition of “vulnerable” provided) [68].

#### 3.3.1 Mortality

Three studies of mortality were identified [59, 69, 76]. In the two instances where children in care were compared to children not in care, children in care had higher mortality rates. Siefert et al. compared statewide mortality rates in 1980–1989 for children aged less than one year in Michigan, USA [76]. Mean mortality rates for children in care were 7.2 per 1,000 children in first care placement compared to 3.7 per 1,000 live births for all children (rate ratio = 1.97; note the denominator for the latter rate includes children that later entered care but these children are a small enough proportion of the population as to not introduce a large error in the rate estimate). The subgroup of African American children in care had yet higher rates with a mortality rate of 9.0 per 1,000 children in first placement (rate ratio = 2.46 compared to all children). The reasons for this, e.g., the increased burden of deprivation on minority ethnic groups, were not explored. Mortality rates were also higher in the county containing the city of Detroit which is relatively more deprived than the state average (rate ratio = 2.50 compared to all children state-wide). The respective mortality rates for children in care and not in care in this county were 9.1 per 1,000 children in their first placement and 4.5 per 1,000 live births, respectively. Caution is required when interpreting the above rates as mortality was rare; over the ten-year study period there were 61 deaths among children in care from a population of 8,597. Where a cause of death was known (n = 53), Sudden Infant Death Syndrome was the most common cause of death for children in care with 28 deaths (52.8%) attributed to it. This was followed by congenital anomalies (28.3%, n = 15).

Chaiyachati et al. also reported mortality rates for children in care to be substantially greater than children not in care using population-level administrative data in the USA [59]. Mortality rates for children in care aged 1–4 years were 50.7 per 100,000 person-years (95% CI: 47.8, 53.6) compared to 27.5 per 100,000 person-years (27.3 to 27.7) for children not in care at the time of death (rate ratio = 1.84; 95% CI: 1.74, 1.95) after adjusting for sex, ethnicity, and race in negative binomial regression models.

Lee et al. examined systemic factors associated with the foster care system in the USA and their relation to mortality rates [69]. Mortality rates in children in care aged 1–4 years were not significantly different from rates in the reference group (children in care aged 15–17 years). More generally, mortality rates did not vary substantially with age except for increased rates in children living in care in the first year of life. No comparisons were made with children not in care.

#### 3.3.2 Physical development

Physical development was considered to include height, weight, and head circumference. When combined, these outcomes were the most common health outcome with 12 of the included studies reporting one of these measurements [49, 51, 57, 60, 62, 63, 66, 72, 75, 77, 78, 83]. Although a consensus definition of failure to thrive (also known as faltering growth) is lacking [85], failure to thrive was considered as a separate outcome as it was often reported independently from height, weight, and ratios of these measurements. Failure to thrive was mentioned in four studies but criteria for this diagnosis were poorly defined in these studies, if defined at all [63, 66, 81, 84].

Of the eight studies that measured children’s height [49, 57, 63, 66, 72, 77, 78, 83], almost all found that care-experienced children were, on average, small for their age. In studies which used population percentiles, all but one [63] found disproportionately high numbers of children below the 5th percentile for age. Estimates in the remaining studies varied from 11% to 38.9% [49, 66, 77, 78, 83]. Halfon also noted that 36.2% (n = 67 of 185) of children aged 0–4 years in their care-experienced sample were under the 25th percentile for height [66]. Other work reported and evaluated heights using standardised *z* -scores. These scores allow a measurement such as height or weight to be reported as a relative non-dimensional value, normalised by the relevant mean value and standard deviation. For example, a *z* -score of -1 means that the measurement is one standard deviation below the mean which always lies at *z* = 0. In the present context, where sets of measurements of children of different ages are aggregated, this allows the relative height or weight of children of different ages to be properly compared since comparison of the absolute measurements would not be meaningful. Bel et al. found that 45.5% (n = 20 of 44) of children aged under two years were more than two standard deviations below mean height-for-age upon entry to care [57]. The mean *z* -score for height was -1.77. In children aged 2–8 years, 13.2% (n = 5 of 36) were more than two standard deviations below mean height-for-age. The *z* -score for height was -0.65. Miller et al. similarly used standardised scores and reported a mean *z* -score of -1.45 for height in a sample of 127 care-experienced children [72].

Ten studies measured children’s weight [49, 57, 60, 62, 63, 66, 72, 75, 78, 83]. As with height, there were disproportionately high numbers of care-experienced children below the 5th percentile for their age. Chernoff et al. found 15.4% (n = 61 of 408) of children aged 0–2 years in this range [49]. Halfon et al. reported a similar figure of 16.8% (n = 31 of 185) for children aged 0–4 years [66]. This proportion was higher among the subgroup of children aged under one year at 20.8% (n = 15 of 72). Wyatt et al. [83] used standardised scores referenced to normative population distributions to assess weight. The median standardised *z* -score for weight-for-age for the sample was -0.16. This contrasted with a positive median standardised score for weight-for-height of +0.30. This was similar to the finding of Dubowitz et al. that only 9% (n ≈ 12 of 137) of care-experienced children aged 0–5 years were in or below the 10th percentile of weight-for-height [63]. Considering all included studies which used standardised *z* -scores demonstrates a pattern of poor development in young children in care. Of the 82 children aged 0–8 years in Bel et al.’s sample, 17.1% (n = 14) were more than two standard deviations below the population mean height for age and sex [57]. The mean *z* -scores for height were -0.72 for children aged under two years and -0.05 for children aged 2–8 years. A mean *z* -score for weight of -1.59 standard deviations was noted by Miller et al. in a sample of 127 care-experienced children [72].

Obesity was less frequently examined than being underweight but was reported in four studies [60, 62, 63, 75]. Croft et al. reported 9.4% (n ≈ 3 of 32) of the subgroup of children aged 2–5 years were obese or overweight [60]. A slightly larger proportion was noted by Dhindsa et al. in this age group (33.3%, n = 3 of 9) [62]. However, both studies were limited by very small sample sizes. A larger study of children in the USA indicated that obesity and being overweight were more prevalent in these groups with 27.7% (n = 23 of 83) of the sample being overweight or obese and 14.5% being obese [75]. This contrasts with the results of an older study conducted in the USA reporting the rate of obesity at 8% of children (n ≈ 11 of 137) in 1992 [63].

Head circumference was measured in five studies [49, 51, 57, 66, 72] and there were typically higher numbers of children with low measurements than expected based on normative population distributions. Using population-derived percentiles, Chernoff et al. reported 18.8% (n ≈ 73 of 408) of children under the 5th percentile for head circumference. Halfon et al. reported 11.9% (n = 22 of 185) of children aged 0–4 years under the 5th percentile and a higher proportion of 19.4% (n = 14 of 72) in the 0–12 months subgroup. Other studies reported results using standardised scores. One noted that 22.7% (n = 10 of 44) children aged under two years had head circumferences which were two or more standard deviations smaller than expected for their age [57]. Another study noted a mean *z* -score of -1.38 SD [72]. In contrast to this finding, Hansen et al. did not find an over-representation in the lowest percentiles for care-experienced children, with 16.3% (n = 13 of 80) of the sample lying in or below the 25th percentile and only 2.5% (n = 2 of 80) lying below the 5th percentile [51].

The lack of a consensus definition of failure to thrive was reflected in the four studies which included this outcome [12, 63, 66, 81]. Dubowitz et al. reported 9% of children (n ≈ 12 of 137) as being in or under the 10th percentile for weight-for-height and defined this as failure to thrive [63]. The remaining studies provided no definition of the condition. Högberg et al.’s population-level study of electronic health records observed that 0.9% (n = 70 of 7,455) of children with an episode of care in the first year of life had been diagnosed with failure to thrive during that period [12]. Halfon et al. reported 10.3% (n = 19 of 185) of their sample with failure to thrive [66]. A higher prevalence of 15.0% (n = 15 of 100) was reported in a study of children aged under three having their health reviewed in a clinic for children in foster care [81]. This relatively high estimate is likely to be due to the clinical setting which over-represents children with more complex needs, even when medical conditions are not the primary reason for children entering care.

Using thresholds for percentiles or standardised scores will fail to identify children that have previously developed well enough to surpass these boundaries but whose growth has subsequently faltered. To explore this issue, Wyatt et al. tracked physical development between entry to care and one year later in 45 children [83]. Development was measured by standardised scores accounting for sex and age. Height and weight scores improved over the study period from -0.21 and -0.16 to -0.02 and 0.35 for height and weight, respectively. The authors also grouped children into three categories depending on the change in their standardised height score. Categories were created using the following thresholds of changes to *z* -scores (Δ*z*): Δ*z* ≥ 0.25 = increase; 0.25 ≥ Δ*z* ≥ −0.25 = neutral; Δ*z* ≤ −0.25 = decrease. The majority of children fell into the neutral (35.6%, n = 16 of 45) or increase (46.7%, n = 21 of 45) categories. This suggests that placement in care had positive effects on children’s physical development as signalled by the improvements in height. The positive effects were felt by all children regardless of baseline status: no correlation was found between baseline height and change in height, and no differences by sex were observed in any outcome.

A second longitudinal study of height and weight also used a one-year follow-up period after entry to care [57]. The results were consistent with those above, where standardised scores for height and weight improved by a significant amount. In children aged under two years upon entry to care, the proportion of those with standardised scores for a height of greater than two standard deviations below normative population average reduced from 45.5% (n = 20 of 44) to 31.8% (n = 14 of 44), with mean *z* -score increasing slightly from -1.77 to -1.51. The respective values for weight were 20.5% (n = 9 of 44) and 13.6% (n = 6 of 44), with a change in mean *z* -score of -1.00 to -0.72.

#### 3.3.3 Dental health

Dental health was a common theme in the included studies with seven papers reporting on outcomes such as dental caries and tooth fracture [17, 33, 52, 55, 63, 78, 80]. The proportion of children with dental caries varied widely across studies. One of the three dental-specific studies identified caries in 57.6% (n = 38 of 66) of children aged under seven years sent for dental health review following entry to care [52]. Notably, 31.8% of the children (n = 21 of 66) had decay in four or more teeth. The rigour of the dental inspection, combined with the lenient inclusion criterion of counting any presence of decay regardless of severity, may account for this high estimate. Regardless, the prevalence of caries was approximately twice that found in an approximately age-matched comparison group that had no experience of care. A perhaps more striking result of this study comes from a comparison of the mean numbers of teeth affected in each group. Children in care had a mean of 3.8 decayed teeth (SD = 0.73) compared to 0.42 (SD = 1.20) in the comparison group.

A second dental-specific study used population-level administrative data to compare outcomes of universal dental reviews for children in care with children with no history of care involvement in Scotland [17]. Among the children in care undergoing the review at age 5, 23.4% (n = 209 of 894) required urgent dental treatment. This was significantly more common than for children not in care with an odds ratio of 2.06 (95% CI: 1.76, 2.42) after adjusting for age, sex, and area-level deprivation. When considering both urgent and non-urgent dental needs 66.6% (n = 595 of 894) of looked after children required further treatment. Such treatment needs remained significantly more common in children in care compared to children not in care (adjusted odds ratio = 2.65; 95% CI: 2.30, 3.05). The proportion of children requiring treatment following the dental review was lower in children in foster care (59%, n ≈ 140 of 237) than children living at home (68%, n ≈ 255 of 375), in kinship care (71%, n ≈ 196 of 276), and residential care (67%, n ≈ 4 of 6).

Studies that used general health reviews found a lower prevalence of dental caries. Swire et al. found caries that was severe enough to warrant referral to a dental specialist in 21% of the 3–5 year olds in their sample (the size of this subgroup was not reported but the overall sample size of children aged 3–15 years was n = 473) [78]. Another study reported an overall prevalence of caries of 13.2% (n = 204 of 1,452) and did not observe significant variation in this proportion by placement type [33] . Dubowitz et al. reported that 15% (n ≈ 21 of 137) of children aged under six years required treatment or referral for caries and other types of dental needs were identified in 7% (n ≈ 10 of 137) of children (children may have appeared in either or both categories) [63]. Takayama et al. only identified caries in 4.0% (sample size reported as ≤ 309) [80]. A small case-control study pairing 25 children in care with 25 age and gender matched children not in care found, unusually, a slightly higher proportion of the latter had tooth fracture or discolouration [55]. However, the difference in proportion (16.0%, n = 4, compared to 28.0%, n = 7) was not statistically meaningful.

#### 3.3.4 Respiratory conditions

Four studies reported on respiratory problems, predominately asthma [33, 63, 66]. For example, one study reported that over 90% of the 19.5% (n = 36 of 185) of children identified with a respiratory problem had asthma [66]. This was described by the authors as “a prevalence rate about three times the national average [observed in all children, regardless of care status]” [66, p.390]. Similarly, another study reported 22.6% (n = 348 of 1,542) of the sample to have an upper respiratory problem with these problems being constituted of “primarily asthma” [33, p.180]. This echoed an earlier estimate from another study which identified asthma in 26.3% (n = 36 of 137) of children [63]. By contrast, Takayama et al. observed asthma in just 3.6% (n ≈ 13 of 361) of children but found upper respiratory conditions (categorised separately from asthma) in 27.4% (n ≈ 99 of 361) of their sample [80]. The difference in estimates may be attributable to the lack of consensus on diagnostic criteria for asthma and the resulting variation in guidelines, especially for preschool children [86].

#### 3.3.5 Haematology and cardiovascular

Anaemia was a commonly assessed haematological condition and was reported in five studies [33, 56, 63, 66, 80]. Lead poisoning was considered by one study [65]. The prevalence of anaemia was reported by Dubowitz et al. at 8.8% (n ≈ 12 of 137) [63]. Similar values were reported in two later studies: 10.0% (sample size reported as ≤ 309) by Takayama et al. [80] and 7.4% (n = 114 of 1,542) by Leslie et al. [33]. In contrast, Halfon et al. identified a higher rate of 22.2% (n = 41 of 185) of anaemic children in their sample [66]. A large and more recent study in the USA reported anaemia in 3.1% of children (n = 11 of 13,323) aged six months or over but under two years [56]. This proportion increased to 3.9% (n = 610 of 15,607) in children aged two years or over but under five years. High blood levels of lead were found by Greiner et al. in 2.7% (n ≈ 17 of 640) of children undergoing health reviews [65]. However, the authors noted that the equipment used to measure these levels were later found to be underestimating lead concentrations. The reported prevalence was therefore likely to be an underestimate. None of these studies included comparator groups to provide comparable figures for children without care experience.

Non-specific cardiovascular conditions were mentioned by two studies [33, 66]. Halfon et al. noted a prevalence of 4.9% (n = 9 of 185). A slightly lower figure of 2.3% (n = 36 of 1,542) was reported by Leslie et al. [33].

#### 3.3.6 Immunisation

Five studies reported on immunisation rates [49, 63, 67, 73, 79]. Care-experienced children were generally less likely to have received full age-appropriate immunisations compared to children without care experience. However, some studies noted that a lack of recorded immunisations could be attributed to missing health records rather than immunisations not having been administered. For example, Dubowitz et al. noted that 41.6% (n ≈ 57 of 137) of the sample of children aged under six were classed as having an “uncertain” immunisation status [63]. Similarly, an earlier study reported that substantial proportions of “preschool” children had no record of being immunised against mumps (68%), measles (36%), rubella (43%), polio (19%), and diphtheria, tetanus and pertussis (23%) [79]. (The sample size and definition of “preschool” children was not reported but around 307 children from the overall sample were under 6 years.) However, the authors were unable to establish what proportion of these children had never received the vaccine as opposed to this being attributable to missing health records. Chernoff et al. reported 30.8% (n = 194 of 630) of children aged under six years had not received full age-appropriate immunisations but did not comment on record missingness [49].

Two studies used designs which partially mitigated the issue of missing health records. The first used a large comparator group of children without care experience (n = 23,919) and found children in care (n = 62) had lower rates for all immunisation types: diphtheria, tetanus, polio (primary) – 90.3% (n = 56 of 62) compared to 96.9% (n = 23,197 of 23,919); diphtheria, tetanus, polio (booster) – 32.2% (n = 20 of 62) compared to 43.4% (n = 10,383 of 23,919); pertussis - 67.7% (n = 42 of 62) compared to 89.8% (n = 21,482 of 23,919); measles, mumps, and rubella - 85.4% (n = 52 of 62) compared to 94.7% (22,672 of 23,919); and haemophilus influenzae type b - 46.7% (n = 29 of 62) compared to 70.7% (n = 16,907 of 23,919) [73]. The second study compared immunisation rates following the introduction of the meningococcal C vaccine in the UK [67]. Administration of this vaccine was more likely to have been recorded on a specific database compared to other universally-offered childhood vaccines. Aggregated over a non-representative convenience sample of county-level health authorities, children in care had a risk ratio of 1.79 of not receiving the vaccine compared to children not in care. However, there was variation between county-level health boards and in some instances the children in care were more likely to have received the vaccine.

#### 3.3.7 Infectious disease

Infectious diseases were reported in three studies [65, 66, 80]. Of children newly placed in foster care who underwent health screening at a specialised clinic in California 19.5% (n = 36 of 185) were reported to have an infectious disease or be affected by a parasite (no detailed information about disease or parasite types was provided) [66]. Skin tests for tuberculosis were carried out in two other studies in the USA (where vaccination for this disease is not routinely administered) and no positive results were returned [65, 80].

#### 3.3.8 Vision and hearing

Vision was reported on by five studies [33, 61, 63, 80, 82]. An early study of ophthalmology outcomes in a sample of children in foster care in New York City, USA, estimated that 24% (n = 15 of 62) of the children aged under two years, and 30% (n = 44 of 148) of children aged 2–6 years, had a problem with their vision [82]. In both age groups, hyperopia (farsightedness) was the most common problem and affected 12.4% (n = 26 of 210) of children aged 0–6 years. In the remaining four studies not specifically focused on vision, there was a lower prevalence of vision problems with estimates ranging from 3.3%–16%. The heterogeneity of assessment methods between studies limit the conclusions which can be drawn from these results.

Hearing was assessed in four studies, all of which examined hearing as part of a broader health review or database review [33, 61, 63, 80]. The prevalence of any hearing issues ranged from 0–6%. However, two of the studies did not assess hearing in younger children as they were not able to engage with the tests in the required manner: Dubowitz et al. only tested children aged over three years [63] and Takayama et al. only tested children aged over two years [80].

#### 3.3.9 Neurological conditions

Neurological conditions were mentioned in six studies [12, 33, 64, 66, 74, 80]. The study of psychotropic medication by Ferguson et al. found that 8.7% (n = 8 of 92) of children in foster care aged under six years had received any prescription [64]. However, only one child received medication to treat an issue which was not behavioural (an anti-hypertensive for treatment of migraine). A review of psychotropic medication use by Radel et al. reported children in care were around twice as likely to have any prescription than children not in care (∼6.5% versus ∼3.0%) [74]. This study is notable for the large sample size of 719,908 children in care and a comparator sample of 31,473,608 children eligible for other forms of subsidised social healthcare in the USA. However, these overall proportions included mental health related medications which are beyond the scope of this study. Yet, when looking across relevant classes of psychotropics, children in care were still more likely to be using antipsychotics (1.06% versus 0.45%), anticonvulsants (0.65% versus 0.22%) antiparkinsonian (0.02% versus *<*0.01%), and medications that target the central nervous system (0.05% versus 0.01%). Two studies that used health reviews found low prevalence of nervous system disorders (2.2%, n ≈ 8 of 361 [80] and 2.5%, n = 38 of 1,542 [33]) but another health review study reported a substantially higher figure (32.4%, n = 60 of 185) [66]. The latter is likely an overestimate as the sample was comprised of children actively referred to a specialist clinic for children in care. Seizures were reported in 0.9% (n = 70 of 7,455) of Swedish children with any recorded care episode aged less than one year by Högberg et al. [12].

#### 3.3.10 Injuries and accidents

Injuries and accidents were mentioned in four studies [12, 18, 66, 80]. Injury, burns, or marks of abuse were relatively uncommon in the two studies which carried out health reviews upon entry to care. Takayama et al. noted that 5.8% (n ≈ 21 of 361) of children had an injury and 4.4% had “marks of abuse” (n ≈ 16; children may have had both outcomes) [80]. Halfon et al. found evidence of injury in only 1.1% (n = 2 of 185) of children [66].

Högberg et al. used population-level administrative health data in Sweden to examine injuries in relation to abuse during the first year of life for children with any episodes of care [12]. A review of medical diagnoses received by these infants generated the following prevalence estimates: head injury or other craniocerebral condition - 4.4% (n = 328 of 7,455); bone fracture - 1.8% (n = 131 of 7,455); burns *<*0.1% (n = 6 of 7,455).

Bramlett et al. used data from a large national telephone survey in the USA to compare high-level health outcomes between adopted children and children living with biological parents [18]. After adjustment for demographic variables that were significantly different between these groups, adopted children were less likely to have sustained an injury requiring hospital treatment in the past year (7.4% and 9.8%; N.B. these proportions arise from multivariable models adjusted for relevant covariates so it is not appropriate to report count data here).

#### 3.3.11 Motor control

Four studies considered fine and gross motor control studies [61, 66, 70, 77]. Simms reported gross motor control problems in 23.9% (n = 27 of 113) of children and fine motor control problems were in 29.2% (n = 33 of 113). Halfon et al. reported gross motor control problems in 34.6% of their sample (n = 64 of 185) and fine motor control problems in 36.2% (n = 67 of 185) [66]. In a cohort of children living in out-of-home care in Australia, Leggat et al. found broadly similar proportions but also that the proportion of children with motor control issues varied depending on the presence of a history of parental substance use [70]. Atypical fine motor control was observed in 22.0% of children (n ≈ 124 of 563) where parental substance use had been present, compared to 30.1% (n ≈ 156 of 519) where parental substance had not been present. Atypical gross motor control was observed in 15.6% of children (n ≈ 89 of 563) where parental substance use had been present, compared to 24.7% (n ≈ 128 of 519) where parental substance had not been present. The authors suggested these findings may reflect that children of parents with substance use problems were more likely to have experienced neglect rather than abuse, and to be placed in kinship care settings where familial relationships were more often maintained. Motor control difficulties were reported in a French study as a proportion of 1,063 diagnoses among 484 children, preventing the calculation of the proportion of affected children [61].

#### 3.3.12 Dermatological

Dermatological conditions appeared in four studies, all of which involved carrying out broad health reviews [33, 63, 66, 80]. In the studies which provided further detail, atopic or seborrhoeic dermatitis were the dominant issues – conditions often linked to poor nappy hygiene. Three of these studies found dermatological issues in 20.5% to 22% of their sample [63, 66, 80] with a fourth reporting a prevalence of 66.5% [33]. In this latter study, prevalence was noted to be broadly the same across care placement types. The exceptionally high prevalence is also likely due to the health review being carried out at the point of entry to care when the acute consequences of neglect would still be present. By contrast, reviews carried out after children had already been in care for a period of time would include children that presumably had received increased healthcare support to treat such conditions [33]. Takayama et al. also observed that the reason for entry to care had a significant influence on the prevalence of skin problems [80]. Children taken into care due to parental neglect were more likely to have a dermatological condition than those subject to parental abuse or caregiver unavailability (e.g., due to incarceration).

#### 3.3.13 Congenital conditions

Congenital conditions were reported in four studies [61, 66, 72, 81]. Halfon et al. reported that 8.6% (n = 16 of 185) of children aged 0–4 years were noted to have a congenital abnormality at screening upon entry to care [66]. A French study reported that congenital conditions comprised 14% of 1,083 medical diagnoses affecting a sample of children in care aged 0–6 years [61]. However, children may have had more than one condition so this figure does not reflect the proportion of children affected. Neither study provided specific details about the distribution of congenital conditions but Down’s syndrome and foetal alcohol disorder syndrome were mentioned as being common.

Two studies explored rates of foetal alcohol syndrome [72, 81]. Miller et al. examined children in a residential care home in Russia, with the authors describing the known excessive ingestion of alcohol in Russia as a motivation for the study. Of the 127 children screened 13.4% (n = 17) had high scores and a further 44.9% (n = 57) had intermediate scores for likelihood of foetal alcohol syndrome. In a more representative general sample in Israel, Tenenbaum et al. reported a prevalence of probable foetal alcohol disorder syndrome of 15.0% (n = 15 of 100) across the care-experienced children that were assessed [81].

#### 3.3.14 Gastrointestinal

A mix of generic conditions were reported by three studies in relation to the gastrointestinal system [33, 61, 66]. A prevalence of 13.5% (n = 25 of 185) was observed among children aged five years or under newly placed in foster care [66]. However, slightly less than half of these cases were infants under one year that displayed symptoms of gastro-oesophageal reflux (i.e., regurgitation of a liquid diet). The same study also noted encopresis (faecal incontinence) in 2.7% (n = 3 of 113) of children aged 1–5 years, an issue noted as being conspicuously absent in a separate study by Dubowitz et al. [63]. Leslie et al. found gastrointestinal issues to be uncommon and reported such problems in only 1.6% (n = 24 of 1,542) of their sample [33]. Gastrointestinal problems comprised 6.7% of 1,083 diagnoses among 484 children in a French sample but, again, the denominator being total diagnoses and not the sample size prevents the calculation of the proportion of affected children [61].

#### 3.3.15 Genitourinary

Genitourinary outcomes were infrequently assessed, with only two studies reporting on them [33, 66]. Leslie et al. reported 1.9% (n = 29 of 1,542) children had a genitourinary issue [33]. A higher prevalence of 7.6% (n = 14 of 185) in children aged 0–4 years was observed by Halfon et al. [66]. Further, enuresis (urinary incontinence) specifically was noted in 6.2% of children (n = 7 of 113) aged 1–5 years. As with encopresis, Dubowitz et al. noted that the lack of children exhibiting enuresis was surprising [63].

#### 3.3.16 Miscellaneous

Several other conditions were reported across the included studies. Halfon’s health reviews of 185 children aged 0–4 years in foster care in Alameda County, USA, found two cases of cancer (1.1%, n = 2 of 185) and 22 cases of recurring ear infection (11.9%) [66]. Leslie et al. reported 2.9% (n = 44 of 1,542) of their sample of children in foster care in San Diego, USA, had a musculoskeletal issue [33] and Dubowitz et al. reported 4% (n ≈ 6 of 137) of their sample of children in kinship care in Maryland, USA, had a referral for specialist orthopaedic treatment [63].

### 3.4 Socioeconomic status and comparator groups

Meaningful assessment of health outcomes in children with experience of care requires comparison with children not in care but living in similar conditions. SES is one of the most important covariates to consider in this respect since living in conditions associated with low SES has substantial negative consequences for health. SES was mentioned in 13 papers (36.1%) as being relevant to child health [12, 17, 18, 49, 51, 56, 58, 63, 69–71, 74, 78]. The authors of these studies recognised that care-experienced children more often come from socioeconomically disadvantaged households and/or neighbourhoods. The areas in which many care placements are situated have relatively greater levels of socioeconomic disadvantage than the median (adoptive households are an exception and tend to be of above-average SES). Despite the authors of these studies recognising the critical importance of adjusting for SES, the methods used often prevented formal adjustment for SES. Only ten studies (27.8% of all included studies) used any kind of comparator of group, of which only five studies (13.9%) were able to implement adjustments when considering results relevant to this study (although some included it as a covariate or control in other analyses not deemed relevant to the present study) [17, 18, 51, 69, 74]. Broadly, children with care experience had poorer health outcomes than children without care experience in the comparator groups, even in those studies that adjusted for SES.

### 3.5 Care setting

Two studies explicitly considered variations in health between children in care while remaining in the parental home, children in kinship care, and children in foster care [17, 33]. Across a range of health conditions grouped into categories broadly reflecting the subsections above, Leslie et al. identified significant differences between care settings for only haematological and neurological conditions. Haematological conditions were found in 4.8% (n = 21 of 438) of children in kinship care compared to 8.4% in both parental (n = 37 of 439) and non-relative care (n = 56 of 665). Neurological conditions were distributed similarly with a prevalence of 0.7% (n = 3 of 438) in kinship care compared to 2.5% (n = 11 of 439) in parental care and 3.6% (n = 24 of 665) in non-relative care. The study of dental health outcomes by McMahon et al. found that children in foster care were less likely to have dental care needs than both children in kinship care and children in formal care in the parental home [17]. Three other studies reported differences by care context but only for results aggregated across children of all ages [61, 71, 75].

### 3.6 Quality assessment

Six studies met the quality criteria specified in the Methods section [17, 18, 51, 59, 68, 74]. Most relied on large administrative datasets or health census data to compare health outcomes of care-experienced children with their peers, except Hansen et al., which reviewed medical records from a single facility [51].

## 4 Discussion

This review collated and synthesised the literature on physical health outcomes affecting young care-experienced children in high-income countries. The health-related conclusions which can be drawn from our results are limited. This is due to the heterogeneity in methods used, the lack of objective criteria for many health conditions, and the paucity of comparator groups in the included studies. These issues result in a lack of context in which to meaningfully interpret the results. Despite this, in the majority of cases where differences were observed between care-experienced children and their peers without care experience, outcomes were poorer in the former group. Future research can be informed by the methodological issues identified by this review.

Height and weight were reported with relative consistency in methodology across studies. These outcomes were among the few where the literature identified by this review unambiguously shows that young children in care are substantially disadvantaged compared to their peers that are not in care. Notable numbers of care-experienced children were small for their age, particularly those aged under two years. Physical growth is typically representative of a child’s development and is a useful proxy for overall physical wellbeing in asymptomatic children. Being small for age is also associated with poorer cognitive ability [87]. Failure to thrive is most often attributable to malnutrition [88], thus treatment of poor physical growth is usually successful in primary healthcare settings [89, 90]. This was reflected by two of the included studies which demonstrated improvements in standardised scores for height and weight after entering care [57, 83]. Longer-term consequences for cognitive and physical development appear to be minor if chronic and cumulative periods of underdevelopment are avoided [91–93]. However, the possible confounding factor of deprivation must be considered. Children living in areas of greater socioeconomic disadvantage are known to have a higher risk of short stature and children in care are more likely to have come from socioeconomically disadvantaged areas [17, 94]. Care-experienced children may benefit from frequent monitoring of height and weight and nutritional interventions that address or avoid periods of poor physical growth, particularly given the association of malnutrition and neglect [95].

Health conditions that are associated with inadequate provision of care were also commonly reported across the included studies. The several well-conducted studies of dental health provided relatively strong evidence that care-experienced children experienced more frequent and more severe dental health issues than children not in care. These results support those found in studies of older care-experiencedchildren [19, 20, 51, 96]. The results for other health outcomes were less definitive. Incomplete immunisation appears to be more common in care-experienced children, although this is partially due to higher rates of missing and incomplete medical records. Skin conditions and anaemia were also mentioned relatively frequently in the included studies. These outcomes can often be prevented through attentive caregiving, such as regular tooth brushing and providing a nutritious diet. However, caregivers can only provide such care when they are not subject to excess levels of stress and have the material resources to do so. Taking dental health as an example, qualitative work has found that dental health in a “vulnerable” population of children was negatively affected when families were undergoing stressful and challenging life events [97]. Additionally, a study of Australian children noted that incomplete immunisation was rarely attributable to parental refusal. In fact, it was often related to stressors such as having a large number of children in the family, having moved home, or children having complex medical needs [98]. The theme that emerges is that abuse and neglect, which care-experienced children have so often been subjected to, are ultimately caused by systemic and structural factors [99]. These factors, such as income inequality, lead to some caregivers facing far greater barriers to their ability to provide high-quality care. Although providing supportive and proactive healthcare services to individuals can mitigate some risk factors, underlying chronic structural factors are far harder to address.

Regarding study methodology, only ten (27.8%) of the 36 included studies used a comparator group [17, 18, 51, 55, 59, 67, 68, 73, 74, 76]. Of these ten, only three (8.3%) used comparator groups either broadly matched for age, sex, and deprivation, or formally adjusted for these variables [17, 18, 51]. Many of the included studies were publications arising as a secondary outcome of health reviews where the primary aim was to identify and treat health problems affecting individual children. As such, it is understandable that matched comparator groups were uncommon. However, children in care disproportionately come from socioeconomically disadvantaged areas and households [17], and deprivation is closely associated with poorer health outcomes [101]. Therefore, it is essential to account for this to present results in a meaningful context. Studies that compared care-experienced children to children without care experience were more likely to acknowledge the importance of socioeconomic factors [100]. This suggests it is feasible for future research to account for the role of deprivation on the health of care-experienced children. Further, types of care settings are not homogeneous and can have a substantive effect on health. For example, children that are adopted often live in households of above average incomes and education levels [102]. Without knowledge of these contextual factors, health policy and practice decisions cannot be evidence-based. By accounting for important covariates and providing context about care settings, future research can increase the impact and utility of its results.

A healthy child is both free of disease and developing appropriately across all aspects of health including cognitive abilities, interpersonal skills, and in many other ways. Past reviews of the health of care-experienced children have shown they experience disproportionately high rates of problems in both physical and non-physical aspects of health [19, 22, 25–28]. Our work provides additional evidence that these children require additional support to mitigate the effects of prior poor treatment on their health. Additionally, some of these reviews identified limitations of the available research similar to those found in our work. Non-representative samples, aggregation of heterogeneous groups of children (e.g., combining age groups and/or care settings), and the lack of comparison groups restrict the specificity of the conclusions which can be drawn from the collated research. The limited available research on the physical health of young care-experienced children restricts the impact that can be had on policy and practice. Ultimately, this restriction prohibits the contribution that current research can make to improving the wellbeing of these children.

### 4.1 Strengths and limitations

The key strength of this study is our use of standardised and systematic methodology that was flexible enough to handle the heterogeneous outcomes and care settings covered by this review. Further, our searches used the heterogeneous terms used in different international legislative areas and cultures to describe children in care. The broad search strategy and the independent screening of all full-text items by two researchers also ensured a large number of relevant studies were captured. The tabulated key results in Table 3 and categorisation of studies by health condition enhance the utility of our work as a quick reference for researchers and practitioners investigating health outcomes in young care-experienced children.

Our review is limited by the exclusion of grey literature and this pragmatic choice may have introduced publication bias [103]. The dominance of studies from a single country (USA) also reduces the generalisability of our results. It is also possible that some children in the included studies were in care due to complex health needs which may be a confounding factor in our synthesis.

### 4.2 Conclusions

This review broadly suggests that young care-experienced children have poorer physical health outcomes than children that have not been in care. However, our results also show the evidence base for this is far from definitive for many health conditions due to limitations of study methodology. Future studies must adjust for demographic differences between care-experienced children and those without by using appropriate comparator groups, characterising the type and length of care when possible, and accounting for the effects of deprivation on health. Although we recognise the difficulty in providing this context, it would greatly increase the value of future research to policymakers and health professionals.

## Supporting information

S2 Table. Database-specific search strategies

S3 Table. Data extraction chart

S1 Table. PRISMA-ScR checklist

## Data Availability

All relevant data are within the manuscript and its Supporting Information files.

## Acknowledgements

The authors would like to thank Valerie Wells of the MRC/CSO Social and Public Health Sciences Unit at the University of Glasgow for her assistance in designing the database search strategies. We would also like to thank Maria Gardani of the School of Health in Social Sciences at the University of Edinburgh for guidance and discussion around review methodology. Finally, we would like to thank Edit Gedeon, Rachel Hamilton and Dylan Lewis of the MRC/CSO Social and Public Health Sciences Unit at the University of Glasgow for their assistance with various parts of this work.

## Supporting information

**S1 Table. PRISMA-ScR checklist.**

**S2 Table. Database-specific search strategies.**

**S3 Table. Data extraction chart.**

