## Supplementary material for "Physical health of care-experienced young children in high-income countries: A scoping review": S2 Table. Database-specific search strategies

Table 1: CINAHL search strategy and result count. Note: Result counts updated on 5th November 2024. Final results restricted to items indexed before 1st October 2024.

| Element | Terms | # results |
|---|---|---|
| S1 | MH(Child, Abandoned OR Child, Adopted OR Child, Foster OR Child, Institutionalized OR Foster Home Care) | 8,629 |
| S2 | TX(care experienced child* OR looked after child* OR lac OR foster* child* OR adop* child* OR orphan*) | 20,062 |
| S3 | TX((“out of home” OR foster* OR “friends and family” OR “kith and kin” OR kinship OR local authorit* OR institution* OR substitute) N1 care) | 14,250 |
| S4 | TX(“care home” OR “group home” OR “child* home”) | 5,372 |
| S5 | TX(“care placement” OR “foster placement”) | 444 |
| S6 | TX(“corporate parent” OR “child* protect*” OR “public care” OR “protective custody” OR “child* welfare” OR “state custody” OR “supported accommodation” OR “supported living”) | 24,423 |
| S7 | MH(“Infant+” OR Child, Preschool) | 407,444 |
| S8 | TX(infan* OR pre-school OR preschool OR baby OR babies OR neonat* OR “under 6” OR “under six” OR “early intervention” OR birth OR “young child*” OR toddler*) | 777,867 |
| S9 | MH(Child Health OR Dental Care for Children OR “Child Health Services+”) | 44,668 |
| S10 | TX(health OR healthcare OR medicat* OR condition* OR patholog* OR feeding OR growth OR thriv* OR stature OR stunting OR immuni* OR infect* OR symptom* OR disease* OR illness* OR “well-being” OR wellbeing OR neurodevelop* OR treatment* OR prescri* OR hospital*) | 6,957,239 |
| S11 | S1 OR S2 OR S3 OR S4 OR S5 OR S6 | 56,142 |
| S12 | S7 OR S8 | 777,867 |
| S13 | S9 OR S10 | 6,960,244 |
| S14 | S11 AND S12 AND S13 | 14,431 |
| S15 | S14 AND EM -20240930 | 14,024 |

Table 2: MEDLINE search strategy and result count. Note: Result counts updated on 5th November 2024. Final results restricted to items indexed before 1st October 2024.

| Element | Terms | # results |
|---|---|---|
| 1 | exp Child, Foster/ | 219 |
| 2 | exp Child, Adopted/ | 175 |
| 3 | exp Foster Home Care/ | 4,019 |
| 4 | ("care experienced child*" OR "looked after child*" OR lac OR "foster* child*" OR "adop* child*" OR orphan*).ab,ti. | 35,102 |
| 5 | (("out of home" OR foster* OR "friends and family" OR "kith and kin" OR kinship OR local authorit* OR institution* OR substitute) adj1 care).ab,ti. | 13,073 |
| 6 | ("care home" OR "group home" OR "child* home").ab,ti. | 5,321 |
| 7 | ("care placement" OR "foster placement").ab,ti. | 499 |
| 8 | ("corporate parent" OR "child* protect*" OR "public care" OR "protective custody" OR "child* welfare" OR "state custody" OR "supported accommodation" OR "supported living").ab,ti. | 8,859 |
| 9 | (infan* OR pre-school OR preschool OR baby OR babies OR neonat* OR "under 6" OR "under six" OR "early intervention" OR birth OR "young child*" OR toddler*).ab,ti. | 1,127,475 |
| 10 | exp Child, Preschool/ | 1,017,050 |
| 11 | (health OR healthcare OR medicat* OR condition* OR patholog* OR feeding OR growth OR thriv* OR stature OR stunting OR immuni* OR infect* OR symptom* OR disease* OR illness* OR "well-being" OR wellbeing OR neurodevelop* OR treatment* OR prescri* OR hospital*).ab,ti. | 16,867,658 |
| 12 | exp Child Development/ | 68,703 |
| 13 | 1 OR 2 OR 3 OR 4 OR 5 OR 6 OR 7 OR 8 | 62,311 |
| 14 | 9 OR 10 | 197,549 |
| 15 | 11 OR 12 | 16,908,761 |
| 16 | 13 AND 14 AND 15 | 6,342 |
| 17 | limit 16 to dt=19000101-20240930 | 6,287 |

Table 3: Web of Science search strategy and result count. Note: Date filter applied to all queries in Web of Science database's graphical user interface to restrict results to those indexed before 1st October 2024.

| Element | Terms | # results |
|---|---|---|
| 1 | TI=("care experienced child*" OR "looked after child*" OR lac OR "foster* child*" OR "adop* child*" OR orphan*) OR AB=("care experienced child*" OR "looked after child*" OR lac OR "foster* child*" OR "adop* child*" OR orphan*) | 54,536 |
| 2 | TI=(("out of home" OR foster* OR "friends and family" OR "kith and kin" OR kinship OR "local authorit*" OR institution* OR substitute) NEAR/1 care) OR AB=(("out of home" OR foster* OR "friends and family" OR "kith and kin" OR kinship OR "local authorit*" OR institution* OR substitute) NEAR/1 care) | 22,702 |
| 3 | TI=("care home" OR "group home" OR "child* home") OR AB=("care home" OR "group home" OR "child* home") | 5,213 |
| 4 | TI=("care placement" OR "foster placement") OR AB=("care placement" OR "foster placement") | 726 |
| 5 | TI=("corporate parent" OR "child* protect*" OR "public care" OR "protective custody" OR "child* welfare" OR "state custody" OR "supported accommodation" OR "supported living") OR AB=("corporate parent" OR "child* protect*" OR "public care" OR "protective custody" OR "child* welfare" OR "state custody" OR "supported accommodation" OR "supported living") | 19,189 |
| 6 | TI=(infan* OR pre-school OR preschool OR baby OR babies OR neonat* OR "under 6" OR "under six" OR "early intervention" OR birth OR "young child*" OR toddler*) OR AB=(infan* OR pre-school OR preschool OR baby OR babies OR neonat* OR "under 6" OR "under six" OR "early intervention" OR birth OR "young child*" OR toddler*) | 1,313,970 |
| 7 | TI=(health OR healthcare OR medicat* OR condition* OR patholog* OR feeding OR growth OR thriv* OR stature OR stunting OR immuni* OR infect* OR symptom* OR disease* OR illness* OR "well-being" OR wellbeing OR neurodevelop* OR treatment* OR prescri* OR hospital*) OR AB=(health OR healthcare OR medicat* OR condition* OR patholog* OR feeding OR growth OR thriv* OR stature OR stunting OR immuni* OR infect* OR symptom* OR disease* OR illness* OR "well-being" OR wellbeing OR neurodevelop* OR treatment* OR prescri* OR hospital*) | 24,535,826 |
| 8 | #5 OR #4 OR #3 OR #2 OR #1 | 97,679 |
| 9 | #6 AND #7 AND #8 | 3,910 |
